# Refugee and asylum seekers’ engagement with health services during pregnancy: A rapid review

**DOI:** 10.1101/2023.11.07.23298208

**Authors:** Jennifer Green, Jane Herbert, Leissa Pitts, Nyari Garakasha, Lisa Gaye Smithers

## Abstract

The objective of this work was to summarise and describe antenatal care experiences of people from refugee and asylum seeker backgrounds living in high-income countries with universal healthcare. Academic articles from six databases and grey literature from selected government websites were systematically searched for English-language articles published 2012-2022. Articles describing perinatal care of people from refugee and asylum seeker backgrounds from the service user and service providers perspective were eligible for inclusion. A thematic synthesis of included articles was undertaken with study quality assessed using Critical Appraisal Sills Program tools.

Of the 37 included articles, there were seven qualitative, 15 quantitative, two mixed-methods studies and 13 reviews. Articles were conducted in Australia (57%), Canada (11%), and the remainder from Europe. Three recurring themes of communication, sociocultural and health system factors were described as barriers or challenges to antenatal care experiences but also presented opportunities for improving care. Many issues around antenatal care experiences for people from refugee and asylum seeker backgrounds remain the same as those identified over ten years ago. To improve antenatal care for people from refugee and asylum seeker backgrounds, health services could implement a range of strategies that support communication, sociocultural experiences and system-related issues.

## Introduction

Antenatal care is essential for protecting and enhancing the perinatal health and wellbeing of parents and newborns. Even in high-income countries, people from refugee and asylum seeker backgrounds are at an increased risk of poor perinatal outcomes than the wider population.(1, 2) For example, refugees and asylum seekers experience more stillbirth (1), emergency caesarean sections,(3) and adverse neonatal outcomes(4) than the general population. While the reasons for such disparities in outcomes are likely to be varied and complex, it is known that people from a refugee and asylum seekers background have lower engagement with health services when pregnant,(5) even when antenatal care is universal.(6) Lower engagement may, in part, be due to the challenges of navigating health care when settling in a new country or because health services are not meeting the needs of people with a refugee or asylum seeker background. Thus, other services and supports may be needed to improve engagement with antenatal care by pregnant refugee and asylum seekers.

For all health services, culturally appropriate and culturally safe practices should be embedded in routine practice.(7) Australian clinical care recommendations have focused on the use of interpreters and, where possible, multicultural health workers.(7) In 2020, 35% of all births in Australia were by people who had been born in another country.(8) Despite ongoing changes to the patterns of refugee arrivals,(9) it has been many years since the Australian recommendations for antenatal care of refugee and asylum seekers have been updated. Thus, we sought to explore the recent experiences of antenatal care provision for refugee and asylum seekers, in health care settings similar to Australia. Applying the ‘population, concept, context’ framework, the relevant population are therefore people with refugee or asylum seeker backgrounds, the concept is the experience of antenatal care and the context is countries with universal health care.

## Materials and methods

This rapid review was conducted in collaboration with the Multicultural and Refugee Health Service of the Illawarra Shoalhaven Local Health District in New South Wales, Australia. The review had an eight-week timeline and intended to inform health service planning. Protocol registration was not undertaken due to the short timeframe. The parameters for the review were discussed between academics and key health service staff (LP, NG) to ensure the review met the needs of the service. The review was conducted systematically, in accordance with procedures for rapid reviews.(10, 11) The review is reported according to the Preferred Reporting Items for Systematic Reviews and Meta-analyses.

### Eligibility

Studies were included if they described the antenatal health care experiences of refugee or asylum seekers in developed countries that provided universal health care. Eligibility was open to systematic, umbrella, scoping or rapid reviews of relevant evidence, as well as empirical studies. Studies published from 2012 onwards were included to capture contemporaneous experiences. Studies were excluded if they did not distinguish the experience of refugee and/or asylum seekers from other immigrants, or if they were not published in English.

Where possible, we applied the UN definition for refugees as persons having a well-founded fear of persecution based on specific grounds and as asylum seekers as someone seeking refugee status. However, we acknowledge that some studies may not define refugee and asylum seeker status or their definition could differ from the internationally-accepted one.

### Information sources

We searched databases that covered medical, nursing and midwifery, public health, psychology and social sciences, which included PubMed, CINAHL, Web of science, PsycINFO, Medline and Scopus. The date of the final database search was August 2022. Grey literature searches included Australian clinical practice guidelines and websites for nursing and midwifery, and for obstetrics.

### Searches

The full search strategy for each database is available from the authors and hits associated with each database are included in the Online Supplementary Appendix A. The searches involved a combination of relevant population-level terms (e.g. refugee* or asylum seeker*), contact with service provision (health or care) and pregnancy-related terms (perinatal, pregnancy, early postpartum, prenatal, antenatal or gestation).

### Screening

Database searches were combined in Endnote software (Version X9). Duplicates were removed using the Endnote function along with reviewing titles in alphabetical order. The final search was uploaded to Rayyan software (online version) for blind review by two authors (JG and LGS). Disagreements were resolved through discussion with a third researcher (JH) or health service staff (NG).

### Data extraction

Information on each study was extracted systematically using a bespoke form that was pilot tested and included the following information: Citation information (e.g. author/s, year, title), type of sample (e.g. refugee, asylum seeker), country/region of research and country of origin for study sample, study aims, study design and data collection technique (e.g. focus group), sample size, outcomes and findings. Data were extracted independently by JG, NG and LS. Discrepancies were resolved through discussion with a third researcher (JH).

### Critical appraisal

Study quality was assessed to describe the literature in this field. The quality of the included studies were evaluated with Critical Appraisal Skills Programme (CASP) tools.(12) Quality assessments of the academic literature were conducted independently by JH and LS.

### Synthesis of results

An adaptation of Thomas and Harden’s method of thematic synthesis was undertaken to summarise the results of the included articles.(13) The synthesis involved a familiarisation phase of reading and re-reading each article, coding or grouping similar results to develop descriptive themes, and reflection in conjunction with health service staff to summarise the final analytical themes. The quality assessments were incorporated into the synthesis through emphasising higher quality studies in the narrative.

## Results

The searches yielded 1392 articles from which 37 full text articles were included (**Fig 1**).

**Fig 1.**
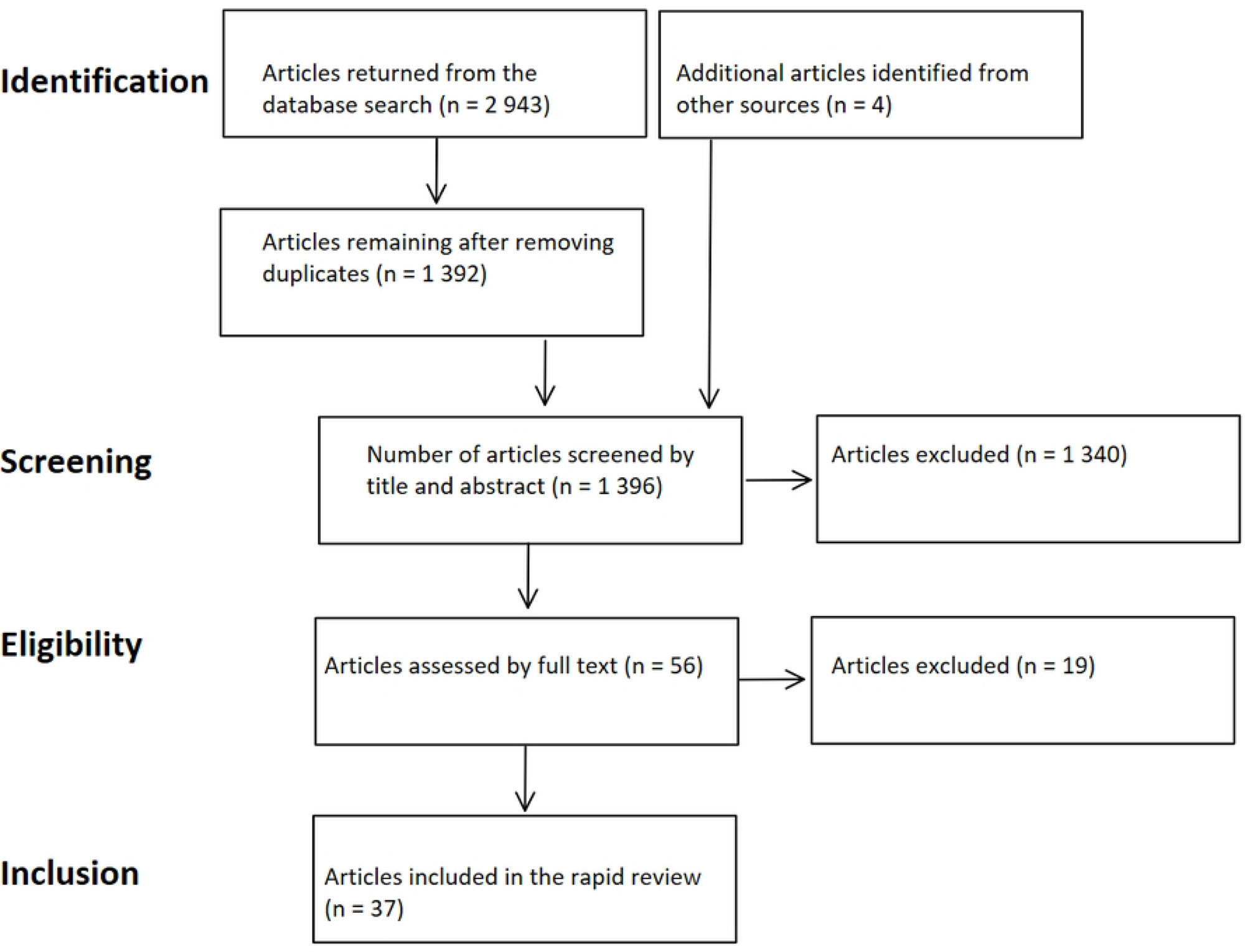
PRISMA flow chart.

**Table 1** describes characteristics of the included studies. These comprised 37 empirical articles of quantitative (*k*=7 (19%)), qualitative (*k*=15 (41%)) and mixed method (*k*=2 (5%)) research, and a further 13 articles that reported on multiple studies (scoping review (*k*=1 (3%)), non-systematic reviews (*k*=3 (8%)) and systematic reviews (*k*=8 (22%) and one systematic review for a national perinatal practice guideline). Most studies were conducted in Australia (21/37 (57%)) with articles also from Canada (4/37 (11%)), mixed countries 4/37 (11%), United Kingdom (3/37 (8%), Finland (2/37 (5%)), Germany (2/37 (5%)), and one each (3%) from Ireland and Sweden. Two articles (5%) involved interviews solely with health service or community workers, 30 (81%) with refugee and asylum seekers, and five (14%) involving both. Thirteen (35%) studies included refugee or asylum seekers from mixed countries of origin, 18 (49%) did not specify their country of origin and the remainder 6 (16%) were of African and/or Middle-Eastern origin. The sample size in quantitative studies ranged from 179(5) to over 34,000 refugees,(3) with qualitative studies ranging from 6(14) to 198.(15) Three recurring themes were reported across articles: communication, sociocultural and health systems factors.

**Table 1:** Characteristics of included studies (*k* = 37) organised by whether it incorporates multiple studies or one empirical article.

| First author (year) | Country of research | Sample ($k$ articles; $n$ participants) | Country of refugee population | Study design | Research information | Main findings |
| --- | --- | --- | --- | --- | --- | --- |
| **Articles that synthesise multiple empirical studies** |  |  |  |  |  |  |
| Balaam et al. (2022) | Europe | Refugee and asylum seeker<br>$k = 16$ | NR | Systematic review | Peer reviewed and grey literature<br>Database inception to 2020<br>English language<br>Qualitative studies | Paraprofessional health workers, community/peer support and less formal interactions valued. Cultural and language barriers resulted in isolation, loneliness, and lack of knowledge about the host country. |
| Behboudi-Gandevani et al. (2022) | Mixed | Migrants from conflict zone countries<br>$k = 40$ | Afghanistan, Eritrea, Ethiopia, Iraq, Kosovo Nigeria, Pakistan, Somalia, Sudan, Syria,, Ukraine, and Yemen | Systematic review | Peer-reviewed<br>English language<br>From 2000 to 2020<br>Quantitative research. | Risk for small gestational age, stillbirth and perinatal morbidity higher amongst immigrants from conflict zones. |
| Billett et al. (2022) | Australia | Migrant and refugee backgrounds residing in Australia<br>$k = 27$ | NR | Systematic review | Academic articles and grey literature<br>Language NR<br>Inception to 2020<br>Qualitative research | Specialist services provided staff knowledge and cultural understanding, flexibility in appointment times, were designed with community participation, bicultural workers and interpreters and continuity of care. Trust between service users and medical staff highly valued. |
| Giscombe et al. (2020) | United Kingdom | Refugee and asylum seeker | NR | Systematic review | Academic and grey literature<br>English only | Both pre and post migration have mental health impacts. |

| First author (year) | Country of research | Sample (*k* articles; *n* participants) | Country of refugee population | Study design | Research information | Main findings |
| --- | --- | --- | --- | --- | --- | --- |
|  |  | *k* = 8 |  |  | 2007-2017<br>Predominantly quantitative research | Social supports are a protective factor for mental health outcomes. |
| Heslehurst et al. (2018) | United Kingdom | Refugee and asylum seeker<br>*k* = 29 | NR | Systematic review (of systematic reviews) | Academic research<br>English articles<br>2007 to 2017<br>Quantitative and qualitative research | Structural, social, personal and cultural barrier to access and use of care exist amongst migrant women. Relationship with healthcare providers and sensitivity to cultural difference noted as positive factors. |
| Iliadou et al. (2019) | Europe | Migrant and refugee<br>*k* = 63 | NR | Literature review | Peer-reviewed and grey literature<br>English articles<br>2002–2017<br>Qualitative research | Major topics for mental health of pregnant immigrants and refugees: Prevalence and risk factors for antenatal mental disorders, assessment of mental disorders, healthcare professionals' training, and mental health interventions. Training in cultural competency is a requirement for effective healthcare, particularly for mental health. |
| Kasper et al. (2022) | Germany | Refugee, asylum seeker and migrant<br>*k* = 16 | NR | Systematic review | Academic articles<br>English or German<br>1990 to 2019<br>Qualitative research | Collaboration between services, strong relationships, time, individualised care and continuity of care were all important factors. |
| Khanlou (2017) | Canada | Migrant and refugee living in Canada<br>*k* = 126 | NR | Scoping review | Peer-reviewed empirical articles and systematic reviews<br>English | Language, cultural differences and knowledge of health system, |
|  |  |  |  |  | 2000 to 2016<br>Qualitative, quantitative and mixed methods | intersecting social experiences were barriers. Poorer perinatal health for refugee and key migrant groups. Country of origin is a strong determinant. |
| Leppalla et al. (2019) | Sweden, Norway, Finland | Refugee<br>*n*=198 women from 10 qualitative studies | 29 different countries | Systematic review | Peer-reviewed research<br>English<br>2013 to 2018<br>Qualitative research | Diminished negotiation power, sense of insecurity and care related discrimination were all barriers for humanitarian migrants. |
| Pangas et al. (2019) | Australia | Refugee<br>*k* = 25 | NR | Meta-ethnographic review | Ethnographic research<br>English<br>2000 to 2017<br>Qualitative | Maintenance of strong cultural identity was important. Difficulty in living between two cultures and negotiating new systems. Impact of trauma. |
| Rogers et al. (2020) | Australia | Migrant and refugee<br>*k* = 17 | NR | Systematic scoping review | Academic and grey literature<br>English<br>2008 to 2019<br>Qualitative, quantitative and mixed methods | having bilingual/bicultural workers, group antenatal care and specialised clinics, culturally responsive care, continuity of care, effective communication, psychosocial and practical support, support to navigate systems, and flexibility in access to services, were all valued by participants. |
| Coe (2021) | Australia | Refugees in Australia<br>*k* = 7 | Afghanistan, Sudan and Myanmar | Literature review | Peer reviewed articles<br>English<br>2010 to 2020<br>Qualitative research | Proximity to service, specialised services (culturally sensitive and continuity of care), social supports, cultural identity and strong relationship with staff were valued features. Access to appropriate translators. |
| Tait (2013) | Australia | Refugee<br>*k* = NR | NR | Non-systematic review | Article sources, dates, languages and data types NR | Importance of professional (not family) translators. Support for cultural training of staff. Awareness needed of pre-existing health issues. |
| Department of Health (2020) | Australia | Migrant and refugee women<br>*k* = NR | NR | N/A, practice guidelines | Details NR.<br>Last updated 2014 | First antenatal visit should have an accredited health interpreter. Multicultural health workers should be involved where possible. Recommends an individualised approach, and working to build trust with patient and explain what services are available. Consider external factors that impact access (e.g. transport). People from refugee backgrounds may face a range of external access challenges. |
| **Empirical research** |  |  |  |  |  |  |
| Markey et al. (2022) | Ireland | Migrants<br>*n* = 52 | NR | Participatory health research design | Perinatal mental health<br>3 online World café sessions (circa 2020)<br>Snowball sampling to capture diverse participation by migrants, health service and community workers | Collaborative whole system approach, cultural sensitivity, trauma informed care, relationships, trained translators and multi- |
|  |  |  |  |  | Thematic analysis<br>Qualitative | language resources, and opportunities to form social connections all valued |
| Leppala et al. (2022) | Finland | Migrants from conflict zone countries<br>n = 3155 (refugee), n = 93600 (Finish born) | NR | Cross-sectional study of health records | Prenatal care<br>Birth registry data 2015-2016<br>Migrants vs Finnish-born women<br>Adjusted logistic regression analysis<br>Quantitative | Migrants from conflict zones less likely to access available care, would access care later, and were less likely to attend the recommended number of check-ups |
| Snow et al. (2021) | Australia | Refugee<br>n = 100 refugees, 100 Australian- born | NR | Review of hospital records | Psychosocial risk and perinatal mental health<br>Consecutive electronic records of refugee vs Australian-born women from July 2015 to April 2016<br>Analysis by Fishers Exact test<br>Quantitative | Refugee participants less likely to report prior mental health issues and financial concerns. Had fewer social supports at antenatal assessment. |
| Henry et al. (2020) | Germany | Refugee<br>n = 12 | Iraq, Syria, Palestine | Phenomenological design | Access & experience of health care and childbirth of Arabic-speaking women<br>Recruited via personal contacts, social workers, waiting room, referrals by refugees<br>Semi-structured interviews Aug-Sept 2017<br>Content analysis using Levesque's access model<br>Qualitative | Strong social support systems, language skills and literacy, continuity of care all led to positive outcomes for participants. Cultural differences in understanding healthcare, and language and access to translators were barriers. |
| Riggs et al. (2020) | Australia | Refugee<br>n = 30 (16 women, 14 men) and an 34 health professionals | Afghanistan | Participatory design | Access to health information<br>Snowball sampling beginning with community consultation (dates NR)<br>Semi-structured interviews and focus groups<br>Thematic analysis<br>Qualitative | Interpreters valued. Access to interpreters (particularly during birth) was a challenge. Participants unsure of role of health professional beyond direct healthcare. |
| Bitar & Oscarsson (2020) | Sweden | Mostly refugee<br>n = 10 | Mixed (Arabic speakers) | Feasibility study | Communication using a tablet<br>Purposive sampling from antenatal clinics<br>Phone interviews Feb 2018 – Mar 2019 in Arabic translated to Swedish<br>Content analysis<br>Qualitative | Evaluation of using health information app. Overall positive feedback from service users for accessing accurate and consistent information about healthcare. Not for more complex questions. |
| Yelland et al. (2020) | Australia | Refugee<br>n = 26 210 (2 740 refugees) | Mixed, >35 countries | Interrupted time series analysis | Access to (number of) hospital-based antenatal visits<br>Routinely-collected hospital data 2014-2016<br>Refugee background vs Australian-born women<br>Logistic regression analysis<br>Quantitative | Evaluation of systems reform in public hospitals. Improvements to attendance at antenatal visits, but steady decrease in proportion of people having their first hospital visit in the first trimester of pregnancy. |
| Leppalla et al. (2020) | Finland | Service providers only<br>n = 18 | NR | Qualitative content analysis of interview data | Hindrances and facilitators to humanitarian migrant maternity care<br>Recruited via staff email (Nov 2017-Sept 2018)<br>Semi-structured interviews<br>Content analysis using the 3-delays model<br>Qualitative | Access to independent and competent translators, and the option of walk in clinics valued. Language, location and navigating bureaucracy were all barriers. |
| Malebranche et al. (2019) | Canada | Refugee and asylum seeker<br>n = 179 | NR | Cohort study | Antenatal care utilization and perinatal outcomes by government- or privately-sponsored refugees vs asylum seekers<br>Electronic medical record review 2011-2016<br>Unadjusted comparisons of outcomes between groups<br>Quantitative | Asylum seekers had more between arrival and first engagement with the service and were more likely to be recorded to have received inadequate care than refugees. No differences were found between asylum seekers and refugees in terms of obstetric and newborn outcomes. |
| Rees et al. (2019) | Australia | Refugee and conflict-affected<br>n = 1335 (289 self-identified as refugees, 396 from conflict areas, 650 Australian born) | Arabic speaking countries, Sudan and Sri Lanka (Tamil-speaking) | Cross-sectional study | Major depressive disorders among refugee vs Australian-born<br>Consecutive refugee and conflict-affected women recruited by trained personnel at antenatal clinics, with parallel sampling of Australian-born via random computer selection procedure<br>Data collection Jan 2015-Mar 2016 by interview<br>Statistical comparisons of outcomes between groups<br>Quantitative | Refugee status associated with a range of intersecting social challenges (e.g. housing, and psychological intimate partner violence). Higher rates of past trauma. Less likely to have strong social supports. |
| Winn et al. (2018) | Canada | Service providers only<br>n=10 | Mixed, Syria influx | Qualitative thematic analysis of interview data | Health care professionals experience of care for pregnant refugee women<br>Purposive snowballing of staff working at a specialist refugee clinic from 2012-2016<br>Semi-structured interviews<br>Thematic analysis<br>Qualitative | Team-based approach, creative communication, developing rapport, commitment and dedication to refugees and extending hours helped service provision.<br>Language, family, culture, lack resources, complexity of health system, migration journey impacts health, and lack of knowledge were barriers. |
| Willey et al. (2018) | Australia | Service providers only<br>n=26 | NR | Inductive analysis of qualitative focus group data | Service provision to pregnant refugee families<br>Purposive sampling of maternal child health nurses via phone or email during May-Jun 2014<br>Six focus groups and individual questionnaires | Service flexibility, communication between services, cultural sensitivity and building relationships with families all valued. |

| First author (year) | Country of research | Sample (*k* articles; n participants) | Country of refugee population | Study design | Research information | Main findings |
| --- | --- | --- | --- | --- | --- | --- |
|  |  |  |  |  | Thematic analysis<br>Qualitative |  |
| Wanigaratne et al. (2018) | Canada | Refugee<br>n = 34 233 (refugee), n = 243 439 (non-refugee immigrants), n = 615 394 Canadian born | Mixed | Population-based data linkage study | Perinatal outcomes of refugee, non-refugee and Canadian mothers<br>Hospital birth records Apr 2002 - Mar 2014<br>Adjusted logistic regression and generalised estimating equations<br>Quantitative | Refugee status showed small associations with some adverse maternal and perinatal outcomes. Refugees also had higher rates of caesarean section, and moderate preterm birth. more detailed information about experiences is required to determine health needs for this cohort, rather than just refugee status. |
| Yelland et al. (2016) | Australia | Mostly refugee families, hospital and community-based health professionals<br>n = 64 (16 Afghan women, 14 Afghan men, 10 midwives, 5 medical practitioners, 19 community-based health professionals | Afghanistan | Thematic analysis of qualitative interview data | Communication during pregnancy, labour and birth<br>Recruitment of Afghans via advisory group, community groups and their leaders; health professionals via key contacts in organisation<br>In-depth interviews and focus groups conducted 2012-2013<br>Thematic analysis<br>Qualitative | Strong reliance on male partners to act as interpreters. Problematic in some scenarios. Extra time is needed for sessions with interpreters. Need for access to professional interpreters. |
| Riggs et al. (2016) | Australia | Refugee<br>n = 19 | Myanmar | Co-designed program evaluation | Group-based antenatal program evaluation<br>Program participants were invited to participate (dates NR)<br>Two focus groups with semi-structured guide<br>Thematic analysis<br>Qualitative | Peer support model resulted in participants feeling better informed, more confident and better connected. Peer support program helped facilitate continuity of care. |

| First author (year) | Country of research | Sample (*k* articles; *n* participants) | Country of refugee population | Study design | Research information | Main findings |
| --- | --- | --- | --- | --- | --- | --- |
| Owens et al. (2016) | Australia | Refugee and culturally and linguistically diverse migrants<br>n = 12 | NR | Phenomenological study | Community-based antenatal program evaluation<br>Purpose sample of women who participated in program<br>Semi-structured interviews with interpreters Jun-Nov 2014<br>Thematic analysis using social constructionist epistemology<br>Qualitative | Community-based antenatal service that specialised in maternity care for CALD women was able to meet the needs of the service users. Using a social model of care focussed on continuity of care and holistic service delivery. Female doctors preferred. |
| Lephard & Haith-Cooper (2016) | United Kingdom | Asylum seeker<br>n = 6 | Sub-Saharan Africa and Eastern Europe | Hermeneutic phenomenological study | Maternity care experiences<br>Purposive and snowball sampling via flyers in children's centres<br>Semi-structured interviews<br>Thematic analysis<br>Qualitative | Community midwives helped connect some participants to health services and to the community.<br>Barriers include communication, knowledge of the health system, associated costs (e.g. transport) social isolation, not being listened to. Sensitivity around female genital mutilation (FGM) needed. |
| Yelland et al. (2014) | Australia | Refugees and health professionals<br>n = 30 (Afghan men and women), n = 34 (health professionals) | Afghanistan | Participatory research | Refugee families experience of maternity services<br>Purposive recruitment via community advisors, organisations, and key stakeholders (for health professionals)<br>Interviews and focus groups 2012-2013<br>Thematic analysis<br>Qualitative | Community based maternal and child health nurses had more capacity to work past the identified barriers as they had better access to interpreters, more time to build relationships with family, and a better understanding of the family's experiences. Healthcare services often were shown to struggle more with providing the above components of healthcare. |
| Gibson-Helm et al. (2014) | Australia | Refugee *n* = 2173 (*n* = 214 North African non- humanitarian source countries (HSC) group), *n* = 1147 (North African HSC group), *n* = 619 (Middle and East African non-HSC group), *n* = 87 (Middle and East African HSC group), *n* = 61 (West African non-HSC group) and *n* = 45 (West African HSC group) | Africa | Electronic database of medical records | Perinatal outcomes of migrants from Africa with and without refugee background<br>Birth data 2002-2011<br>Analysis by adjusted logistic and linear regression<br>Quantitative | Poorer health outcomes, and higher frequencies of negative health experiences around pregnancy indicate that refugees from African regions face additional barriers compared to African migrants from non-refugee source countries. Socio-economic disadvantage, the need for interpreter, particular health needs around FGM and various health outcomes more common amongst refugees from some source countries compared to others. |
| Renzaho & Oldroyd (2013) | Australia | Migrant *n* = 35 | Afghanistan, Africa, China, Palestine, Lebanon, Syria, Iran, Jordan | Thematic analysis of qualitative interview data | Sociocultural barriers and health needs in pregnancy and postnatal period<br>Participants recruited through playgroups 2010-2012<br>Five focus group discussions with interview schedule and bilingual worker<br>Thematic analysis<br>Qualitative | Importance of social connections and contacts for positive health. Need for service providers to understand diversity in what this looks like. |
| Niner & Kokanovic (2013) | Australia | Refugee and asylum seeker *n* = 15 | Myanmar | Thematic analysis of qualitative interview data | Birthing experience of Karen women in Australia<br>Purposive recruitment via community networks (dates NR) | Past experiences of trauma, lack of knowledge of health system, language and access to translators and |

| First author (year) | Country of research | Sample (k articles; n participants) | Country of refugee population | Study design | Research information | Main findings |
| --- | --- | --- | --- | --- | --- | --- |
|  |  |  |  |  | Interviews in Karen with a bilingual researcher<br>Thematic analysis<br>Qualitative | perceived negligence of staff all impacted participants engagements in Australia. Strong feelings of gratitude, self-reliance and stoicism amongst participants. |
| Stapleton et al. (2013) | Australia | Refugee women (quant) n = 190<br>Other women (quant) n = 4158<br>Refugee women (qual) = 18<br>Staff n = 5<br>Researchers and community workers n = 10<br>Service user survey = 42<br>Staff survey = 147 | Somali, Sudanese, Afghan, Burundi, and Liberia | Mixed methods | Cross cultural use of EPDS<br>Recruitment information NR<br>8 focus groups, surveys by women and staff, medical record audit of refugee and other women; dates NR<br>Thematic analysis of qualitative data; descriptive analysis of quantitative data<br>Quantitative and Qualitative | Cultural sensitivity and impact of cultural differences needs to be acknowledged and catered for (e.g. understanding of collective rather than individualistic cultural norms) |
| Stapleton (2013) | Australia | Refugee women n = 42<br>Other women n = 4158<br>Hospital staff n = 150<br>Interpreters n = 2<br>Community stakeholders n = 5 | Mixed | Mixed methods using data from a program evaluation | Refugee women's experiences of antenatal clinics<br>Refugee women recruited through family and community networks, staff via workplace communications and emails<br>8 focus groups, surveys by women and staff, medical record audit of refugee and other women; collected Jan 2009-May 2010<br>Thematic analysis of qualitative data; descriptive analysis of quantitative data<br>Quantitative and qualitative | Continuity of care, personalised care, and flexibility all highly valued by participants. Staff training and accessible location of service recommended. Acknowledgement that these features often cost more. |

### Communication

Many articles noted language and communication as a barrier to perinatal care(15–31) and identified opportunities to improve communication through translators. Language challenges experienced at booking in particular created difficulties in managing appointments and knowing what services are available.

Studies noted that people from refugee and asylum seeker backgrounds take longer to engage with antenatal services than the general population, access less healthcare overall, and less healthcare than recommended.(6, 32) In addition to difficulties with knowing what services are available or how to access them,(14–16, 18, 25, 26, 28, 33) uncertainty about their residency status meant that people from a refugee or asylum seeker background did not consistently know what they were allowed to access, or whether their access rights may have changed over time.(14, 22, 26) These issues may be overcome by ensuring additional time is provided at the point of initial service engagement and throughout the perinatal period to explain the health care system and what birthing in a local hospital is like.(34) A strong relationship with a healthcare provider supports people from a refugee and asylum seeker in finding information and services they need from pregnancy to infant wellbeing.(21, 25, 35)

Providing information about services and accessibility in a variety of languages and formats was consistently identified as important for reducing the information needed to be covered by translators. However, low literacy among some people from a refugee background meant that written materials remained inaccessible.(19, 20) Access to professional translators helped ensure accurate and confidential information was being communicated between service users and healthcare professionals, although issues of concern included availability of translators,(1) their ability to communicate in the same dialect,(6) and their age and gender appropriateness.(1, 18) Appointments facilitated by translators took longer,(28, 36) but this was not discussed as having been accounted for with longer appointment times.(34, 36) Family members (particularly male partners) often acted as a translator, which was a practice supported by participants.(21) However, when family members acted as translators, health providers expressed concern about the accuracy of the information being translated,(37) whether true informed consent to medical treatments was provided,(38) and privacy when discussing sensitive issues such as domestic or family violence.(39)

### Sociocultural

Common concepts raised in the included studies involved acknowledging culture, providing culturally-sensitive care and avoiding cultural stereotyping. The combination of best-practice care delivered in a culturally-safe manner was highly valued by people from refugee backgrounds.(18, 35, 38) Several studies cited maintenance of a strong cultural identity,(19) and feelings of self-reliance and stoicism(36, 40) among participants. Some reported the feeling of living between two cultures, which was challenging to navigate.(36)

Some articles noted that cultural practices around pregnancy and childbirth were not possible for some people from refugee backgrounds. For example, the involvement of older female family members (for support and guidance) was not possible if those relatives did not live in Australia, which was upsetting.(20) The role of family during pregnancy and birth caused conflict between some participants from refugee backgrounds and their healthcare providers, particularly among less individualised cultures (than, for example, Australia).(18, 19, 36, 41) Health care providers’ acknowledgement of cultural challenges was an important element of person-centred culturally-responsive care,(18, 20) and was facilitated by the formation of strong and safe relationships between service providers and service users, cultural sensitivity and clear communication about the host country health system. When information or services differed from participants expectations, conflicted with their cultural understanding, or were not thoroughly explained in a culturally safe way, it often led to the person refusing or ignoring the advice, or feelings of isolation, sadness and of not being heard.(14, 26, 30, 35) Culturally-responsive care was underscored by ongoing staff training around cultural differences and providing trauma informed care.(19, 33, 34, 42) Barriers were identified around understandings of mental health and stigma in discussing mental health concerns.(25, 43) with staff training considered important for identifying mental health concerns.(42) Avoiding cultural stereotyping or making assumptions about cultural backgrounds or refugee experiences was also important.(17, 25, 26, 31)

Socioeconomic factors were associated with the refugee experiences and also impacted on access to healthcare. Examples include unstable, uncertain or unsuitable housing, transport challenges and costs, and experiences of poverty.(4, 14, 18, 19, 22, 25, 26, 35) Refugee and asylum seekers expressed concern about costs of services (or assumed costs if they were unsure of their entitlements),(14) although this varied by country.(3)

### System

The included articles identified a range of system-level influences such as staff-related factors, services that provide collaborative and holistic care, and offered continuity of care. Refugee and asylum seekers may have a limited understanding of how the health system of their host country functions, the medical model of childbirth and the different role/s different healthcare workers and services. Providing information about these systems may remove barriers and improve trust.(15)

Having staff who understand and act on the needs of people from refugee and asylum seeker backgrounds was identified as essential for optimal and effective healthcare.(14, 17, 24) The included articles discussed training in cultural responsiveness and trauma-informed care (particularly on mental health problems and parenting) as well as providing culturally-sensitive care.(31, 33, 34) Culturally competent care should be individualised rather than generalised to avoid cultural stereotyping of an individuals’ cultural background or refugee experience.(17) Other challenges for healthcare staff include having sufficient knowledge and resources for responding to non-clinical needs such as access to social supports,(17, 29) and responding sensitively to specific issues such as female genital mutilation.(14, 18, 31)

Services that were valued by people from refugee and asylum seeker backgrounds included continuity of care,(17–20, 27, 28, 34, 38) collaborative care,(17, 28, 33) working in multidisciplinary teams,(14, 16, 18) and paraprofessional health advocates. Continuity of care was noted as particularly important for strengthening relationships between healthcare workers and refugees. Often, specialist services were set up well to provide this model of care,(18) largely due to their capacity to provide flexible and more personalised care. This demonstrated a strong commitment to people from refugee and asylum seeker backgrounds and built trust. Collaboration between medical and social services helped people access health services unrelated to their pregnancy and social supports beyond healthcare.(19, 27, 28, 43) Models of care that included social elements were reported to be better equipped at providing for social needs than hospitals. Sometimes these models were created through a co-design approach.(28, 38) Paraprofessionals helped facilitate connection, trust and continuity for people from a refugee background with the health service.(16) Community-based nurses were often thought to have better access to interpreters, more time to build relationships and a better understanding of the family’s experiences.(29)

While a range of services may be available, many services work in silos, which limits collaboration and sharing of expertise and access to services.(33) Continuity of care supported positive relationships between the healthcare provider and people from refugee and asylum seeker backgrounds and improved rapport.(27) A specialist service involving translators or bilingual workers was supportive of provider-patient rapport.(20) Better coordination of care between services, and communicating the specific needs of individuals from refugee backgrounds, may improve care and barriers to access.(17) Collaborative models engaged with community organisations (e.g. playgroups, religious groups and multicultural centres) fostered connections between community and services, and could connect people from refugee and asylum seeker backgrounds in both directions, by acting as referral pathways to better improve service access and by providing culturally-accessible education programs, specialist group clinics and community-based pregnancy support.(35)

Flexible services in accessible locations were highly supported by people from refugee backgrounds as they enabled greater access and engagement with services.(18, 34, 35, 38) Service location mattered as transport to services can be a barrier to access,(34, 35) and services with flexible appointment scheduling were important.(34, 35) Furthermore, services that provided a range of medical and social supports enabled refugees to access health services unrelated to their pregnancy.(19, 27, 28, 43) As refugee and asylum seekers experience social isolation,(18, 43) services that helped connect with people who spoke their language or had shared experiences were valued.(14, 19, 20, 27)

None of the included articles focussed specifically on experiences of racism within the healthcare system, beyond identifying where the healthcare provided was not culturally sensitive. Experiences of discrimination impact upon refugee and asylum seekers trust in host country services.(2, 15, 18, 25, 30) And where mistrust in authority and government occurs in their home countries, negative experiences in the host country can exacerbate refugees lack of trust and result in poorer healthcare.(15)

### Study quality

Assessment of each article’s quality is shown in **Online Supplementary Appendix B Table S2,** as a function of study design. Most studies were rated as medium quality, with clear aims, appropriate methods, and valid data. Potential biases during recruitment or design stages were common, including a lack of detail on how refugee status was defined(5) and the broad use of convenience sampling methods in qualitative studies, such as personal networks,(20, 39) recruiting through a specialist playgroup(40) or referrals received from community members, social workers or health care professionals.(20, 28, 30) In many studies the impact of these pre-existing relationships on design decisions or the generalisability of the results was not given adequate consideration.(14, 27, 29, 30, 33, 37) While most studies reported findings of local relevance, a few exceptions were noted for very small *(n* = 6) interview studies(14) and reviews incorporating papers with migrants from a wide range of conflict zones(1) or host countries with a vastly different healthcare system than Australia (e.g. USA)(38). Older studies were rated as lower quality(30, 31, 34)because they reported less detail on design and analysis decisions, potentially reflecting changes in publishing norms. The grey literature(7) was assessed as low quality due to the rapid review format and presentation style but was of the highest local relevance.

## Discussion

In the current scoping review, we identified 37 articles reporting various aspects of perinatal care for people from refugee and asylum seeker backgrounds. Although articles were published from many different countries, three recurring themes were identified. These included challenges with communication, sociocultural and health care system factors. Research quality was moderate, with most articles reporting well-defined aims, appropriate designs, and being of value. We identified potential biases from pre-existing relationships on recruitment, design decisions and generalizability of the data. Taking a strengths-based perspective, evidence from the three main themes present a range of opportunities to improve antenatal care experiences. Translators and multicultural health workers can support communication. Acknowledging culture, training and delivering culturally-safe and trauma-informed care were important for building trust and maintaining engagement with services. Providing information about the health system and healthcare providers’ roles in the host country may be helpful. Working collaboratively with other health, social and community services and providing continuity of care in convenient locations were highly valued. Although catering to diverse service users can be problematic, improving staff access to resources perhaps through resource sharing across institutes and providing staff professional development are addressable health service factors.

Although a third of the included articles involved reviews or meta-syntheses of individual studies, our review is distinct because we focus on the *experience* of perinatal care rather than specific perinatal outcomes (e.g. postnatal depression, caesarean section) and our target population are refugee and asylum seekers rather than general migrant populations. Despite the different focus, we report consistencies with past reviews. For example, Heslehurst et al conducted a review of systematic reviews and, consistent with the current review, issues raised included communication challenges, a lack of cultural knowledge and sensitivity, cultural stigma and stereotyping, the importance of supportive relation with practitioners, and a desire for integrated care and ability to include cultural practices in the host settings.(25) Reviews that included older literature raised similar concerns such as sociocultural challenges associated with migrating to a Western country,(44) engagement with antenatal care could be improved with culturally-sensitive, non-stigmatising care and collaboration with other (community) agencies.(45) Wikberg & Bondas(46) discussed issues of respect, active listening and socioeconomic disadvantage. Thus, although the literature included in these reviews was ineligible for the current study because they were published over 10 years ago, the consistency with the current review suggests that improving antenatal care for migrant, refugee and asylum seeker people has been slow and many issues remain unaddressed.

Notably, most empirical research targeted refugee and asylum seekers from mixed countries of origin. The implications of this are that the current body of research will reflect more generalised experiences that are not specific to particular cultural groups. This is both a strength and a weakness of the research to date and its ability to inform clinical practice. On one hand, high income countries that offer universal health care must provide perinatal care to service users from diverse backgrounds. Hence the findings of this review are applicable to a similarly ‘general’ group from which it was derived. Yet, the backgrounds, cultural practices and health care needs are sometimes unique to particular cultural groups, which means we have less specific information on how to improve clinical care for such groups.

A challenge with the literature in this area is that there is no consistent definition of refugee and asylum seekers. Many empirical articles and reviews often do not define people from refugee or asylum seeker backgrounds, and there is no standard or agreed way of asking about migration status and refugee experience.(29) which may lead to challenges in providing tailored service responses. Implications for the current review are while participants differ across studies, the similarities in themes across studies suggests this has largely not impacted review findings. Other limitations of this review include the 10-year time horizon, limiting to English language publications and high-income countries with universal health care. These constraints were imposed to ensure the review was contemporary, a manageable size and expense, and able to inform innovations to current practice in an Australian setting. This review is likely to be of use to similar settings, such as the New Zealand, UK and Canadian health services.

## Conclusion

Key themes relating to the experience of antenatal care for people from refugee and asylum seeker backgrounds involve communication, sociocultural and systems-level factors. While elements of these themes are similar to those identified over ten years ago, the evidence synthesised here suggests that strategies are available for health services to address these issues and improve the antenatal care experience for people from refugee and asylum seeker backgrounds.

## Data Availability

All data used for the study is available from the primary research articles.

## Acknowledgements

We would like to acknowledge the health service advice and feedback on our manuscript from Dr Smriti Jaiswal.

## Supporting information

S1. **Search strategy and summary of hits**.

S2. **Quality assessments for quantitative and qualitative studies, and reviews.**

## Supporting information

### Appendix A: Search strategy description and hits

Table S1: Summary of hits for each database

### Appendix B: Quality assessments for quantitative and qualitative studies, and reviews

Table S2: Summary of quality assessments for quantitative studies

Table S3: Summary of quality assessments for qualitative studies

Table S4: Summary of quality assessments for review articles

